# Opening schools and trends in SARS-CoV-2 transmission in European countries

**DOI:** 10.1101/2021.02.26.21252504

**Authors:** Alessandra Buja, Matteo Paganini, Vittorio Cristofori, Tatjana Baldovin, Riccardo Fusinato, Giovanna Boccuzzo, Silvia Cocchio, Silvia Coretti, Vincenzo Rebba, Maria Parpinel

## Abstract

**Background:** It is important to understand the role of schools in the community transmission of SARS-CoV-2, bearing in mind that children and adolescents can spread the infection within families, even when their own symptoms are mild. The aim of this study was to examine the trends of contagion before and after schools reopened across 27 countries in the European Union.

**Methods:** All data on the number of people testing positive for COVID-19 in each European country were collected from 20 days before schools reopened to 45 days afterwards. The Joinpoint regression method was used to detect single change points on the trend of contagion. The Bayesian Information Criterion (BIC) was used for model selection purposes.

**Results:** We calculated 27 linear regression models for the daily case numbers of SARS-CoV-2 infection in the 27 countries from 20 days before schools reopened to 45 days afterwards. A significant increase in the number of daily infections was seen for 21 countries after a change point in the linear regression lines. The change points in different countries varied, ranging from 10 to 42 days after schools reopened, with the majority occurring beyond the 21st day.

**Conclusion:** This study analysed the trend of SARS-CoV-2 transmission before and after schools reopened in Europe. We observed a significant increase in the number of new daily cases in most countries. This issue poses a public health problem that needs to be taken into account in deciding strategies to contain the spread of COVID-19.

## BACKGROUND

A large-scale epidemic of COVID-19 caused by the severe acute respiratory syndrome coronavirus 2 has been occurring in Europe since January 2020 ^1^. To mitigate community transmission of COVID-19, European governments introduced sweeping non-pharmaceutical interventions (NPIs) during the first and second waves of the epidemic. These measures have varied from one country to another, but have generally involved isolating confirmed cases, quarantine for contacts, social distancing, international and national travel restrictions, and the closure of schools and non-essential (or less-essential) shops and services ^2^.

The specific burden of each measure is hard to estimate because they have often been imposed more or less simultaneously, and also because testing policies have changed over time ^3^.

It is important to understand the role of schools in the community transmission of SARS-CoV-2, bearing in mind that children and adolescents can spread the infection within families, even when their symptoms are mild ^4^.

Many countries and media have reported clusters in kindergartens, and in primary and secondary schools ^5-8^. The prevalence of confirmed cases of SARS-CoV-2 infection in schools seems to be influenced mainly by the level of transmission in the local community ^9^. Transmission between children in the same class seems to be rare ^10^, due to the use of face masks, frequent hand washing, social distancing, and aeration. The risk of infection differs for different age groups, however. There is evidence of contagion rates usually being low in early childhood ^11,12^, probably due to children being less susceptible to SARS-CoV-2, while markedly higher rates are seen among adolescents. Attending school involves groups having lessons together in classrooms for several hours, and also other issues unconnected to face-to-face teaching, such as the use of public transport and eating meals, which need to be taken into account. Another important factor concerns social contact, which is known to be greater among adolescents, whose school attendance is often combined with other types of behaviour that could facilitate the spread of viral infections, such as the use of public transport, meeting in bars or similar venues after lessons, and studying together. The contribution of this age group to the transmission of COVID-19 remains unclear, however.

In this study, we examined the trends of contagion before and after schools reopened across the 27 countries of the European Union.

## METHODS

Data on the daily numbers of people testing positive for COVID-19 for each European country were obtained from the Our World in Data (OWID) Coronavirus pandemic website, which is updated daily ^13^. OWID is a project run by the Global Change Data Lab, a registered charity in England and Wales. All data were collected from 20 days before schools reopened in each country to 45 days afterwards in 2020. Spain has not provided data on the number of new cases of infection identified between Saturday and Sunday of each week since January 2020.

The dates when schools restarted in each European country were obtained from documents published by the Education, Audiovisual and Culture Executive Agency (EACEA) as part of its Education and Youth Policy Analysis ^14^, based on national data. As the starting dates varied between different European countries, and sometimes also between different levels of education, and different regions of the same country, we considered the date when most of the schools started in a given country.

The dependent variable was the raw count of new cases of COVID-19 detected each day. The number of days elapsing before or after schools reopened was considered as an independent variable. A Joinpoint regression model with a Grid Search method was adapted to the data grouped by country, based on the assumption of uncorrelated errors, in order to detect a single change point on the trend of contagion. Then, whether the change point was statistically significant or not, two different regression lines were estimated, both as follows:

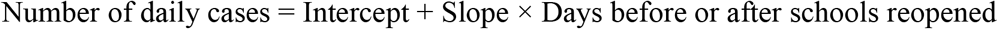

Then only the first line was retained for the whole observation period if the change point was not statistically significant. The Bayesian Information Criterion (BIC) was used for model selection purposes. All reported p-values are two-sided, with a significance threshold of p < 0.05. All statistical analyses were performed using the Joinpoint Regression Program in the SEER*Stat software, version 4.8.0.1.

## RESULTS

We performed 27 linear regression models for the daily numbers of SARS-CoV-2 infections recorded in the 27 European countries from 20 days before schools reopened to 45 days afterwards. A significant increase in the number of daily infections was observed for 21 countries after a change point in the linear regression lines, as shown in *Table 1*. The change points in the various countries ranged from 10 to 42 days after schools reopened, with the majority occurring after the 21^st^ day.

**Table 1:**
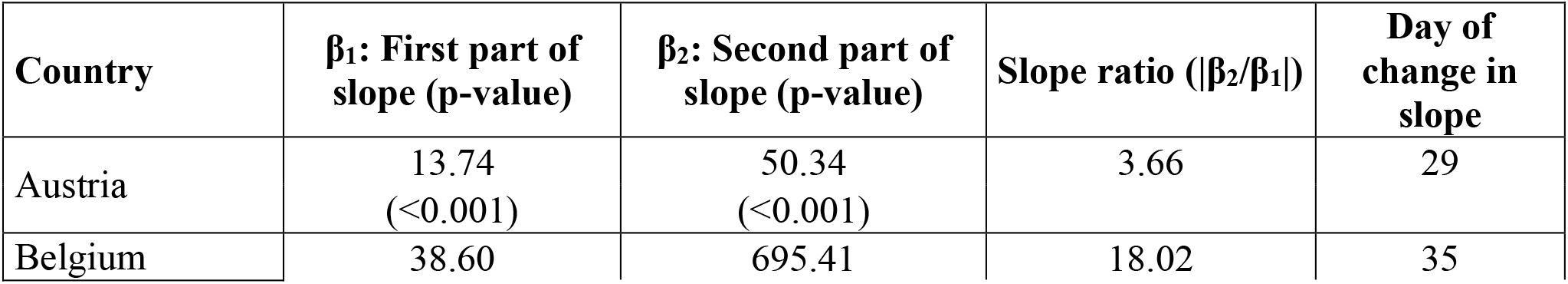
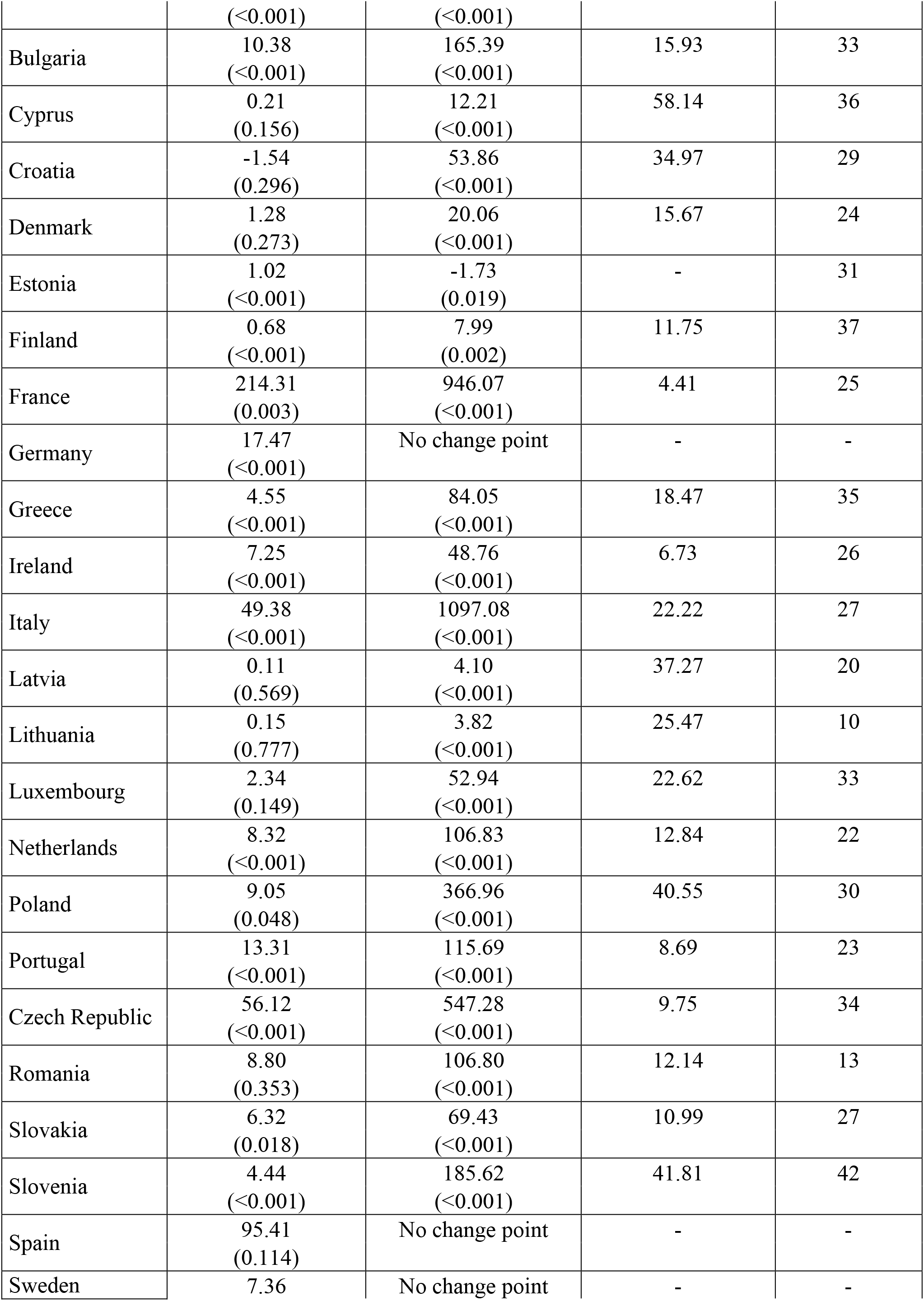
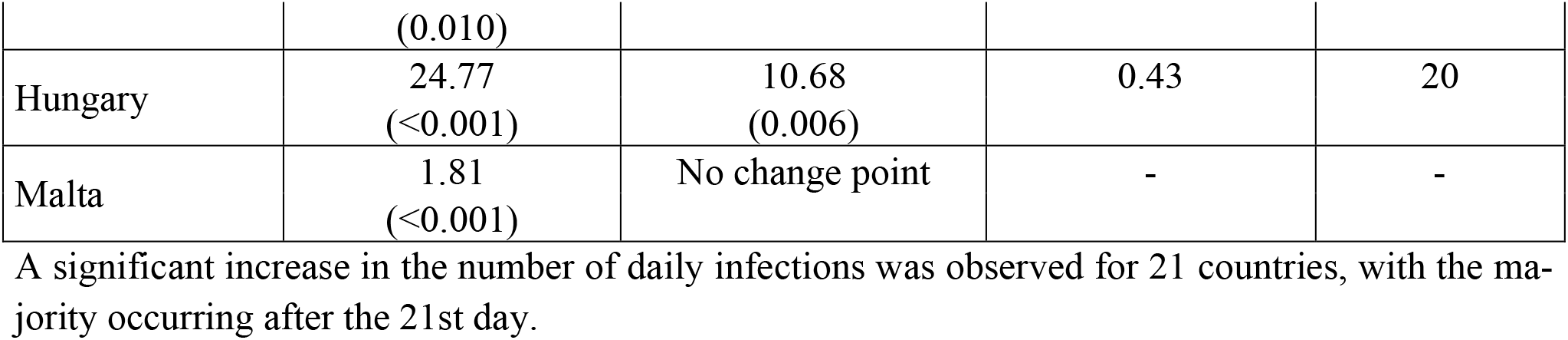
Contagion trends in 27 European countries

In Hungary the trend appeared to slow beyond the change point. Only Estonia showed a decrease in the trend of infections after schools reopened. The numbers of new cases detected each day in Germany, Spain, Sweden and Malta did not seem to increase after schools reopened (no change points were identified).

The slopes of the linear regression lines beyond the change points increased by less than 5 times for Austria and France, between 5 and 10 times for the Czech Republic, Portugal and Ireland, between 10 and 25 times for Luxembourg, Italy, Bulgaria, Belgium, Denmark, Finland, Slovakia, Greece, the Netherlands and Romania, between 25 and 50 times for Lithuania, Poland, Latvia, Slovenia and Croatia, and over 50 times for Cyprus.

*Figure 1* shows the changes in the slope for 8 European countries after schools reopened: Cyprus (β2/β1 58.14); Slovenia (β2/β1 41.81); Poland (β2/β1 40.55); Croatia (β2/β1 34.97); Italy (β2/β1 22.22); Belgium (β2/β1 18.02); Hungary (β2/β1 0.43), the only country where the trend of the slope slackened off beyond the change point; and Estonia, the only country where the trend of the contagion seemed to drop after schools reopened.

**Figure 1:**
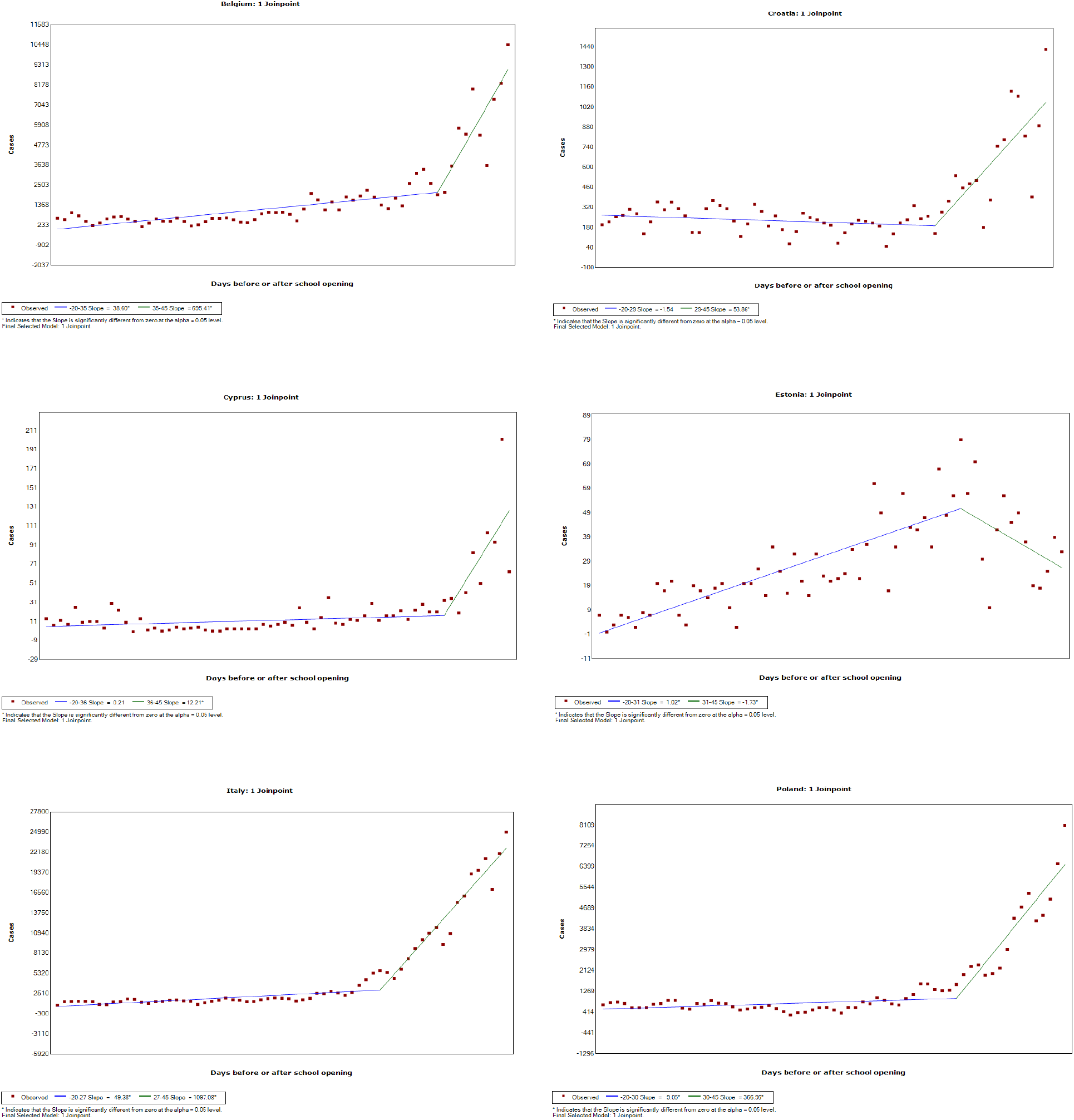
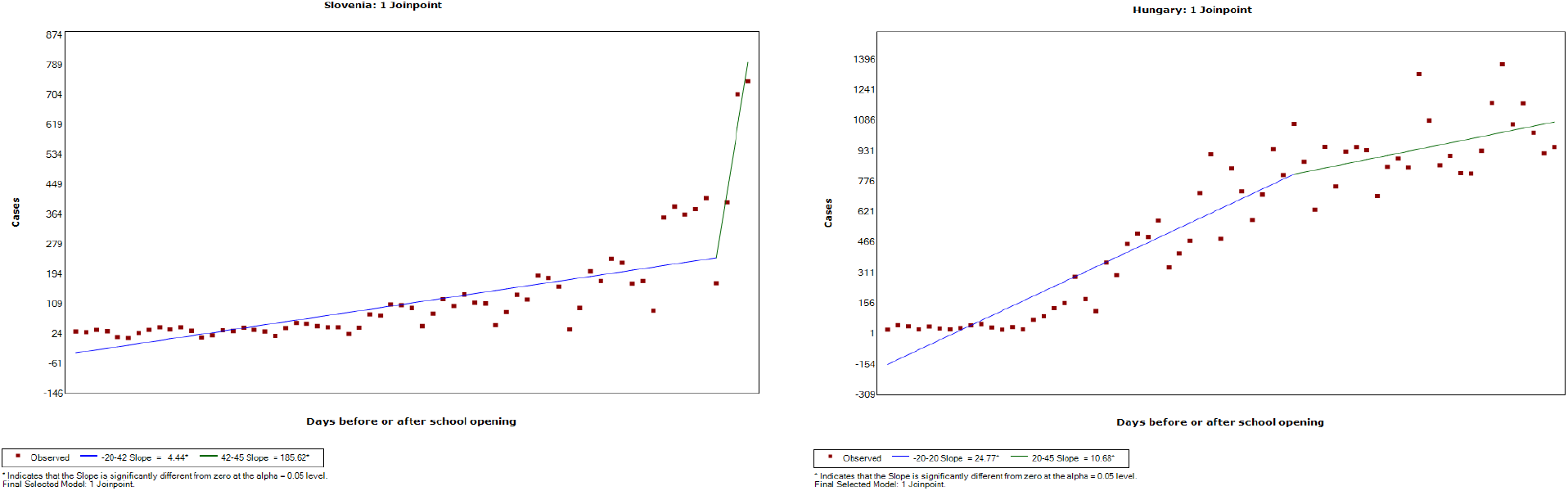
Trend in numbers of new SARS-CoV-2 infections in selected countries over a period of 65 days (20 before and 45 after schools reopened)

## DISCUSSION

This study analysed the trends of COVID-19 transmission before and after schools reopened in Europe. There was a significant increase in the number of new daily infections during the first 45 days after school started for 21 of the 27 EU Member States. The trend of the contagion worsened in almost all European countries after a change point that occurred from 10 to 42 days after schools reopened. This finding indicates that caution is needed in assessing the cost-benefit balance when designing lockdown loosening policies. National school closures to stem the COVID-19 pandemic had been implemented by 107 countries around the world by 18 March 2020, and in 192 countries by the end of April ^2, 15^. The timing of their reopening has varied considerably across and within countries, and for different levels of education ^16^.

A study analysing the relative rates of infection by age group identified higher rates among adolescents and young adults than among children during the first wave of the epidemic in Germany [RR (age group): 0.78 (10-14); 1.14 (15-19); 1.40 (20-24); 1.06 (25-29)] ^17^. Another study conducted to identify sociodemographic factors behind the spread of COVID-19 found that provinces with lower aging indexes had higher rates of contagion, suggesting that younger people could be more responsible for spreading the virus at population level ^18^. Opposite findings emerged for Hungary, however: a recent publication showed that the trend of contagion slowed beyond the change point after schools reopened. New cases of infection may have gone under-diagnosed, however, considering the high rate of positivity among the swabs obtained and the fact that Hungary had the lowest ratio of swabs per 1000 population of all European countries during the period observed ^19^.

Our study revealed no change points after schools reopened for Germany, Malta, Sweden or Spain, although the numbers of new infections were still growing in these countries. In Estonia the rate of infection even seems to have dropped after schools reopened. It is important to bear in mind, however, that we considered only a short-term follow-up (45 days), and it may be that rising case numbers do not become apparent for up to two months after relaxing such measures as school closures ^20^.

Our study has a major limitation that needs to be mentioned: we investigated changes in the slopes of SARS-CoV-2 contagion after schools reopened, but this does not imply a causal inference. “Before and after” analyses typically suffer from several internal validity issues, one of which may concern the effect of the passage of time: changes in an outcome measure might be due to some other influential event(s) occurring in the meantime. In our scenario, for instance, the reopening of sports facilities or seasonal temperature changes (which induce people to engage in different leisure activities) could have affected our outcome variable. In other words, the reopening of sports facilities, greater access to leisure activities, and a more frequent use of public transport may have contributed to the rise in disease transmission rates recorded after schools reopened ^2^.

The management of school closures as a measure to contain the COVID-19 pandemic has represented a major challenge in European countries. Governments have faced a hard trade-off between safeguarding the population’s health and assuring young people’s education. In many cases, the reopening of schools during the pandemic was hotly criticised by the public. However, one-hundredseven countries by March 18, 2020 and 192 countries by April, 2020 implemented national school closures in response to the COVID-19 pandemic ^2, 15^. The duration of school closures, and thus the timing of re-opening, has been very heterogeneous across countries and among school degrees with-in countries ^16^.

In conclusion, our results show that reopening schools correlated with a faster diffusion of SARS-CoV-2 in most EU countries. That said, school closures and, more broadly, the suspension of face- to-face education have negative consequences for students of various ages. As well as interfering with their academic progress, they can give rise to higher dropout rates, higher economic costs for families ^21^, and depression and anxiety in the young ^22^. This means that various decision-making criteria and several different perspectives need to be taken into account when establishing policies relating to pandemic containment measures ^23^.

## Data Availability

All data are from publicly available repositories, as listed in the references section.

